# Neural response to repeated auditory stimuli and its association with early language development in children with Fragile X syndrome

**DOI:** 10.1101/2022.07.05.22277114

**Authors:** Winko W. An, Charles A. Nelson, Carol L. Wilkinson

## Abstract

**Background:** Fragile X syndrome (FXS) is the most prevalent form of inherited intellectual disability and is the most common monogenetic cause of autism. Previous studies have linked the structural and functional alterations in FXS with impaired sensory processing and sensory hypersensitivity, which may hinder the early development of cognitive functions such as language comprehension. In this study, we compared the P1 response in event-related potential (ERP) and its habituation to repeated auditory stimuli in male children (2-7 years old) with and without FXS, and examined their association with clinical measures in these two groups.

**Methods:** We collected high-density electroencephalography (EEG) data in an auditory oddball paradigm from 12 children with FXS and 11 age- and sex-matched typically developing (TD) children. After standardized EEG pre-processing, we conducted a spatial principal component (PC) analysis and identified two major PCs — a frontal PC and a temporal PC. Within each PC, we compared the P1 amplitude and inter-trial phase coherence (ITPC) between the two groups, and performed a series of linear regression analysis to study the association between these EEG measures and several clinical measures, including assessment scores for language development, non-verbal skills, and sensory hypersensitivity.

**Results:** At the temporal PC, both early and late standard stimuli evoked a larger P1 response (p = 0.0037, p<0.0001, respectively) and higher ITPC (p = 0.0402, p = 0.0027) in FXS than in TD. We observed habituation of ITPC in both groups at the frontal PC (p = 0.0149 for FXS; p = 0.0244 for TD). Linear regression analysis showed that within the FXS group reduced frontal P1 response to late standard stimuli and increased habituation were associated with better languages scores. No associations were observed with non-verbal skills or sensory hypersensitivity.

**Conclusion:** We identified P1 amplitude and ITPC in the temporal region as a contrasting EEG phenotype between the FXS and the TD groups. P1 response and habituation in the frontal region may be reflective of the language outcome in male children with FXS. These EEG measures are potential biomarkers for early diagnosis and future language development in patients with FXS.

## 1 Introduction

Fragile X syndrome (FXS) is a genetic condition that affects approximately 1 in 4,000 males and 1 in 8,000 females (Rais et al., 2018). It is the most prevalent form of inherited intellectual disability and is the most common monogenetic cause of autism (Crawford et al., 2001; Hersh et al., 2011). FXS results from an expansion (>200 repeats for full mutation) and hyper-methylation of a CGG trinucleotide repeat in the *FMR1* (Fragile X messenger ribonucleoprotein 1) gene, and individuals with this mutation exhibit developmental and behavioral challenges including delays in learning, speech and language delay, sensory issues, hyperactivity, and anxiety (NICHD, 2021). The long repeat of the CGG sequence prevents the expression of the encoded FMRP protein, which leads to alterations in the development of synapses, including thin and elongated dendritic spines with increased density, and immature synaptic connections, as evidenced by studies with FXS animal models and postmortem studies of FXS individuals (Rudelli et al., 1985; Comery et al., 1997; Hinton et al., 1991; Irwin et al., 2002). It might also prevent activity-based synapse maturation and synaptic pruning, which are essential in developing normal cognitive functions (reviewed in Schneider et al. (2009) and Knoth and Lippé (2012)).

The structural and functional alterations in FXS have been linked with atypical neural processing and arousal modulation problems (Barnea-Goraly et al., 2003). Previous research using rodent models with *FMR1* knockout (KO) mice revealed cortical hyperexcitability due to impaired inhibition and altered neural synchrony (Gonçalves et al., 2013; Zhang et al., 2014). A decreased level of gamma-aminobutyric acid (GABA) receptors and GABAergic input, and increased GABA catabolism were observed in multiple regions in the *FMR1* KO mouse brain (Idrissi et al., 2005; Selby et al., 2007; D’Hulst et al., 2009). This deficit of GABAergic inhibition impacts multiple components of the sensory and cognitive system, including the auditory brainstem (McCullagh et al., 2020), amygdala (Olmos-Serrano et al., 2010), and the auditory cortex (Song et al., 2021), and may underlie the auditory hypersensitivity and auditory processing alterations commonly seen in FXS (Castrén et al., 2003; der Molen et al., 2012a; Schneider et al., 2013; Rotschafer and Razak, 2014). Notably, previous studies in autism have associated auditory processing alterations with language delays (Rincón, 2008; Roberts et al., 2011), a phenotype often shared by the FXS population (Abbeduto et al., 2007; Finestack et al., 2009), which suggests a tight relationship between the auditory response of the brain and language development in individuals with FXS.

In the recent decades, noninvasive neuroimaging techniques like electroencephalography (EEG) have made it possible to track dynamical brain responses without the need of complex neurosurgery, and thus have become a popular tool for studying auditory brain responses. With EEG collected from the widely-used auditory oddball paradigm, previous studies identified multiple components in the event-related potential (ERP) being altered in individuals with FXS relative to the control group. Elevated N1 (Clair et al., 1987; Castrén et al., 2003; der Molen et al., 2012b; Knoth et al., 2014; Ethridge et al., 2016) and P2 (Clair et al., 1987; der Molen et al., 2012b; Knoth et al., 2014; Ethridge et al., 2016) amplitudes have been consistently reported from individuals with FXS. Habituation of N1, defined as the reduced neural response (i.e., the N1 response) to repeated stimulus presentations, was shown to be weaker in FXS compared to age-matched comparison groups (Castrén et al., 2003; der Molen et al., 2012a; Ethridge et al., 2016). Similar to the findings in ERPs, greater response and weaker habituation were also observed in gamma (>30Hz) oscillatory power and inter-trial phase coherence (ITPC) in individuals with FXS (Ethridge et al., 2016). Nonetheless, most of these studies have focused on the adult population; very little is known about the developmental aspect of these neural signatures in FXS. Wunderlich et al. (2006) explored the maturation of auditory ERP in infants and young children, and discovered that the waveform and scalp distribution of auditory ERP change as a function of age. For example, the P1 component, which is prominent in preschoolers and young children, is diminished in adults (Wunderlich et al., 2006; Kuuluvainen et al., 2016). This is possibly the reason why none of the aforementioned studies reported significant findings in P1 response. Additionally, the association between EEG signals and early language development in FXS remains unclear. To our knowledge, only one study by Wilkinson and Nelson (2021) addressed this question, and reported a positive relationship between resting-state gamma power and language scores in male children with FXS. This finding is important from a clinical perspective, because therapeutics (both behavioral and pharmacologic) will ideally be provided near the age of diagnosis (∼3 years of age). More EEG biomarkers that could sensitively, robustly, and timely predict future language outcomes in FXS children are therefore desirable for the implementation of early intervention.

To fill these gaps in research, this study recorded EEG data in a passive auditory oddball paradigm from preschool and school aged boys with or without FXS and collected clinical and behavioral measures. First, we compared the amplitude and short-term habituation of the P1 response and its corresponding ITPC between the two participant groups. We hypothesized that the FXS boys would have greater amplitude and less habituation in these measures than their age-matched typically developing peers. Second, to investigate the clinical relevance of these EEG measures, we examined how they are associated with language development, non-verbal skills, and sensory hypersensitivity.

## 2 Materials and Methods

### 2.1 Participants

A total of 16 children (33 - 78 months old) with Fragile X syndrome (FXS) and 13 age-matched, typically developing (TD) children (33 - 80 months old) were recruited for participation. The uneven number of participants between groups was a result of the COVID-19 interruption. All FXS participants had documented full mutation of the *FMR1* gene. Given that female FXS patients have variable expression of the *FMR1* encoded protein and thus variable phenotypes, they were excluded from this study (n = 1 in the FXS group). Three boys with FXS did not perform or complete the EEG experiment of this study. Additional exclusion criteria across both groups (FXS and TD) included a history of prematurity (<35 weeks gestational age), low birth weight (<2000g), known birth trauma, known genetic disorders (other than FXS), unstable seizure disorder, current use of anticonvulsant medications, and uncorrected hearing or vision problems. Only children from families whose primary language is English (> 50% of the time at home) were included. Some participants were on stable doese of medications (Oxybutin (1 TD); Melatonin (2 FXS); Miralax (1 TD); Sertraline (1 FXS)). More information about the participants can be found in Table. 1.

**Table 1:** Sample characteristics

|  | FXS | TD |  |
| --- | --- | --- | --- |
|  | N = 12 | N = 11 | p value |
| Age, mean in months (SD) | 50.68 (16.66) | 49.03 (12.82) | 0.7914 |
| Maternal Education , n (%) |  |  |  |
| <4-year college degree | 1 (7.69) | 0 (0) |  |
| 4-year college degree | 4 (30.77) | 4 (36.36) |  |
| >4-year college degree | 8 (61.54) | 7 (63.64) |  |
| Paternal Education, n (%) |  |  |  |
| <4-year college degree | 4 (30.77) | 0 (0) |  |
| 4-year college degree | 2 (15.38) | 5 (45.45) |  |
| >4-year college degree | 7 (53.85) | 6 (54.55) |  |
| Household Income, n (%) |  |  |  |
| <\$40,000 | 0 (0) | 0 (0) | |
| \$40-70,000 | 3 (23.08) | 0 (0) | |
| \$70-100,000 | 3 (23.08) | 3 (27.27) | |
| \$100-140,000 | 2 (15.38) | 4 (36.36) | |
| >\$140,000 | 4 (30.77) | 4 (36.36) | |
| Race |  |  |  |
| White | 9 (69.23) | 6 (54.55) |  |
| African American | 0 (0) | 1 (9.10) |  |
| Asian | 1 (7.69) | 1 (9.10) |  |
| Mixed | 1 (7.69) | 3 (27.27) |  |
| Ethnicity, n (%) |  |  |  |
| Hispanic or Latino | 3 (23.08) | 0 (0) |  |
| Clinical Measures, mean (SD) |  |  |  |
| Nonverbal Developmental Quotient | 58.56 (15.56) | 112.49 (16.59) | <0.0001 |
| PLS Auditory Comprehension | 69.54 (15.09) | 118.64 (9.88) | <0.0001 |
| PLS Expressive Communication | 68.77 (17.15) | 122.09 (8.78) | <0.0001 |
| VAS Receptive Language | 9.15 (3.56) | 14.55 (1.51) | <0.0001 |
| VAS Expressive Language | 6.62 (3.50) | 15.55 (1.37) | <0.0001 |
| Sensory Profile | 23.36 (6.58) | 23.56 (5.10) | 0.9437 |

This study was approved by the Institutional Review Board at Boston Children’s Hospital / Harvard Medical School (IRB#P00025493). Written informed consent was obtained from all guardians upon their children’s participation in the study.

### 2.2 EEG collection and experiment design

The EEG was recorded in a dimly lit, sound-attenuated, electrically shielded room. Participants either sat in their caregiver’s lap or sat independently in a chair, high-chair or stroller depending on their preference, and the caregiver was instructed to avoid social interactions or speaking with their child. During the experiment, EEG data were collected using a 128-channel HydroCel Geodesic Sensor Net (Version 1, EGI Inc, Eugene, OR) connected to a DC-coupled amplifier (Net Amps 300, EGI Inc, Eugene, OR) with impedance of all electrodes kept below 100 kΩ. Data were sampled at 1000 Hz with reference to the electrode Cz. A sequence of 800 tones were played with 1000-ms inter-stimulus interval at 70dB from a speaker while the child was watching a silent movie for compliance (Figure. 1). These tones were all 50 ms in duration, and could be either 1000-Hz or 2000-Hz. The 1000-Hz tones, deemed as the more frequent “standard” (STs) stimuli, appeared for 87.5% of the time, while the 2000-Hz tones, deemed as the less frequent “deviant” stimuli, accounted for the rest 12.5%. The number of STs preceding a deviant was either 4, 5, 6, or 7, varying with equal probability.

**Figure 1:**
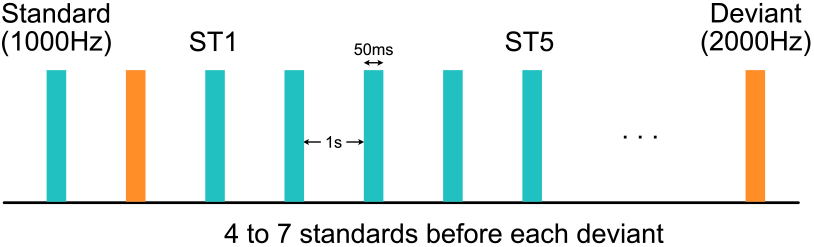
Experimental design of the auditory oddball paradigm. Each standard or deviant stimulus was 50ms in duration. Inter-stimulus interval was 1 second. The first and the fifth standard stimuli after each deviant stimulus were deemed as the “ST1” and the “ST5” conditions, respectively.

### 2.3 EEG pre-processing

Raw EEG data were exported from NetStation (version 4.5, EGI Inc, Eugene, OR) and were batch preprocessed with the Harvard Automated Preprocessing Pipeline for EEG plus Event-Related Software (HAPPE+ER) (Monachino et al., 2021), a MATLAB-based EEG processing pipeline. The processing steps in HAPPE+ER are as follows. Line noise was first removed using CleanLine via a multi-taper regression approach. Signals were then resampled to 250 Hz and low-pass filtered (100 Hz). Bad channels, including those with flat line, residual line noise, and other excessive noise evaluated by a joint probability method, were removed and interpolated. A subsequent wavelet-thresholding artifact removal pipeline (Castellanos and Makarov, 2006) — an algorithm that parses signals into frequency components and identifies artifacts based on the distributions of these components — was implemented to remove noise in the frequency domain. After band-pass filtering (1 – 30 Hz), continuous EEG data were then segmented into epochs between 200 ms before and 500 ms after the onset of each stimulus, and were baseline corrected by the average over the pre-stimulus period (−200 to 0 ms). Only those epochs associated with the first (ST1) and the fifth (ST5) standard tone after each deviant were retained in this study. It should be noted that “ST5” was chosen for analysis for two reasons. First, it is more distant from “ST1” than any earlier standards, and thus is expected to elicit more habituation effects. And second, the number of trials in “ST5” (n = 92) is closer to the number of trials in “ST1” (n = 100) compared to any later standards (n < 65). Epochs with residual artifacts, evaluated by their amplitude and joint probability, were removed by HAPPE+ER. We excluded participants with fewer than 20 trials in either ST1 or ST5 condition — 12 participants in the FXS group and 11 participants in the TD group remained in this analysis after the overall EEG pre-processing. In order to control for the difference in number of trials between ST1 and ST5, we randomly downsampled the condition with more trials to match with the other condition.

### 2.4 EEG analysis

An overview of the EEG analysis is shown in Figure. 2. In brief, a spatial principal component analysis was performed on preprocessed EEG data, and the principal components were applied on the original multi-channel EEG signals as spatial filters to derive PC-transformed time courses. Event-related potential and inter-trial phase coherence measures were calculated from these time courses, which were later studied for their association with clinical measures of different domains.

**Figure 2:**
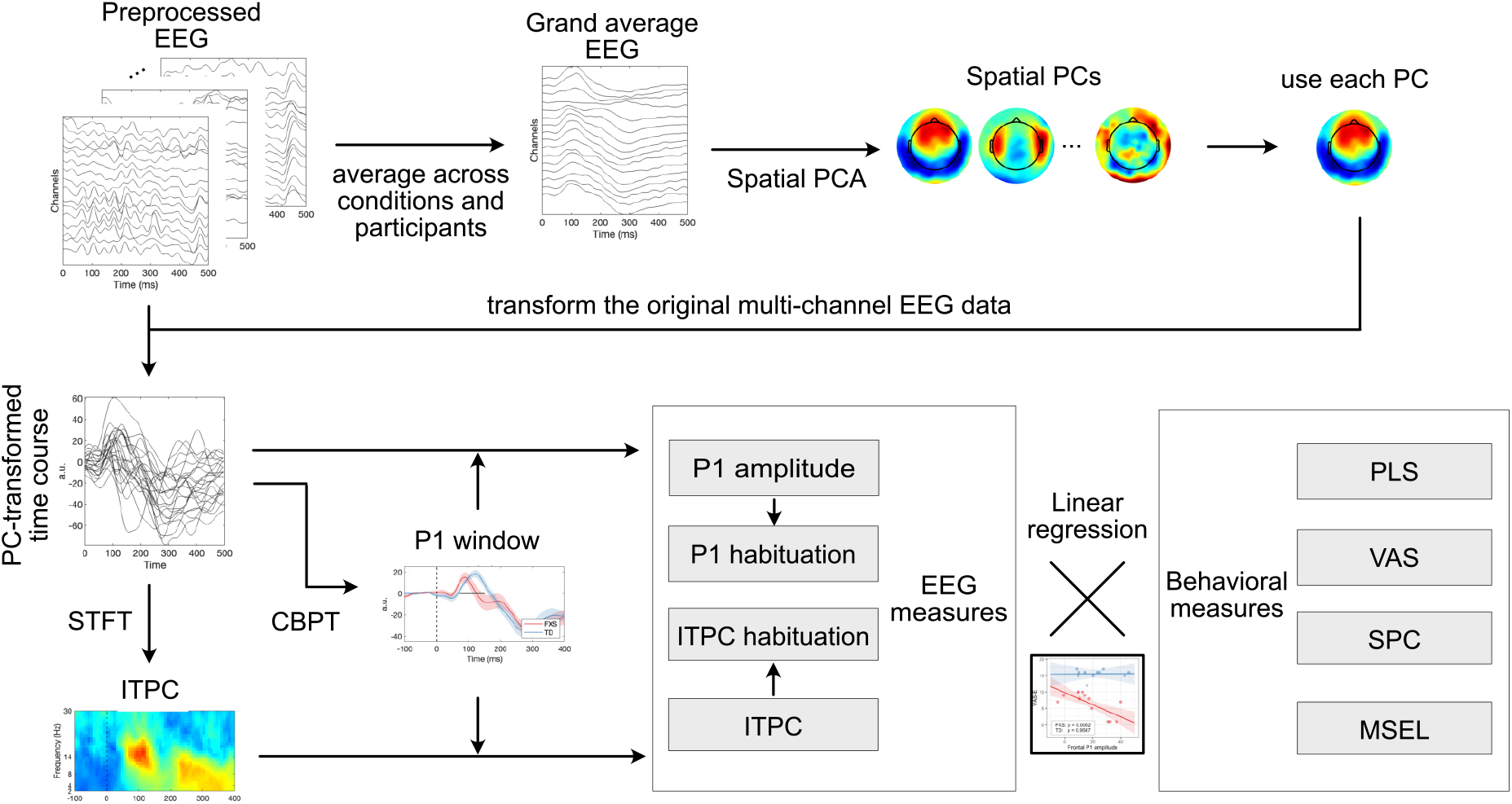
A schematic diagram for all the analyses in this study. For intelligibility, only 16 out of 128 channels are displayed in “Preprocessed EEG” and “Grand averaged EEG”. Each line in “PC-transformed time course” represents the average ERP of an individual. PCA, principal component analysis; a.u., arbitrary unit; STFT, short-time Fourier transform; ITPC, inter-trial phase coherence; CBPT, cluster-based permutation test; PLS, Preschool Language Scales; VAS, Vineland Adaptive Behavior Scales; SPC, Sensory Profile Child; MSEL, Mullen Scales of Early Learning.

#### 2.4.1 Principal Component Analysis

A spatial principal component analysis (PCA) was conducted to identify representative spatial patterns in neural activation following the steps in previous studies (Ethridge et al., 2012, 2015, 2016). EEG data were first averaged across trials, conditions and participants for a grand average EEG matrix, whose dimensions are number of time points by number of channels. A spatial PCA processed each time point as an observation and each channel as a variable, and generated a series of mutually orthogonal principal components (PCs) that could sequentially explain most of the variance in the data. The number of PCs equals the number of time points (i.e., more than 100), and most of them carry a negligible amount of variance explained. In order to determine the number of PCs that should be included for further analysis, we ran a parallel analysis (Franklin et al., 1995) implemented in MATLAB (Shteingart, 2022), which identified two PCs as being significant with more than 95% confidence (Figure. 3). We deemed the one with high weights in the frontal region as the “frontal PC”, and the one with high weights in the temporal regions as the “temporal PC”. All subsequent analyses were performed in these two PCs independently.

**Figure 3:**
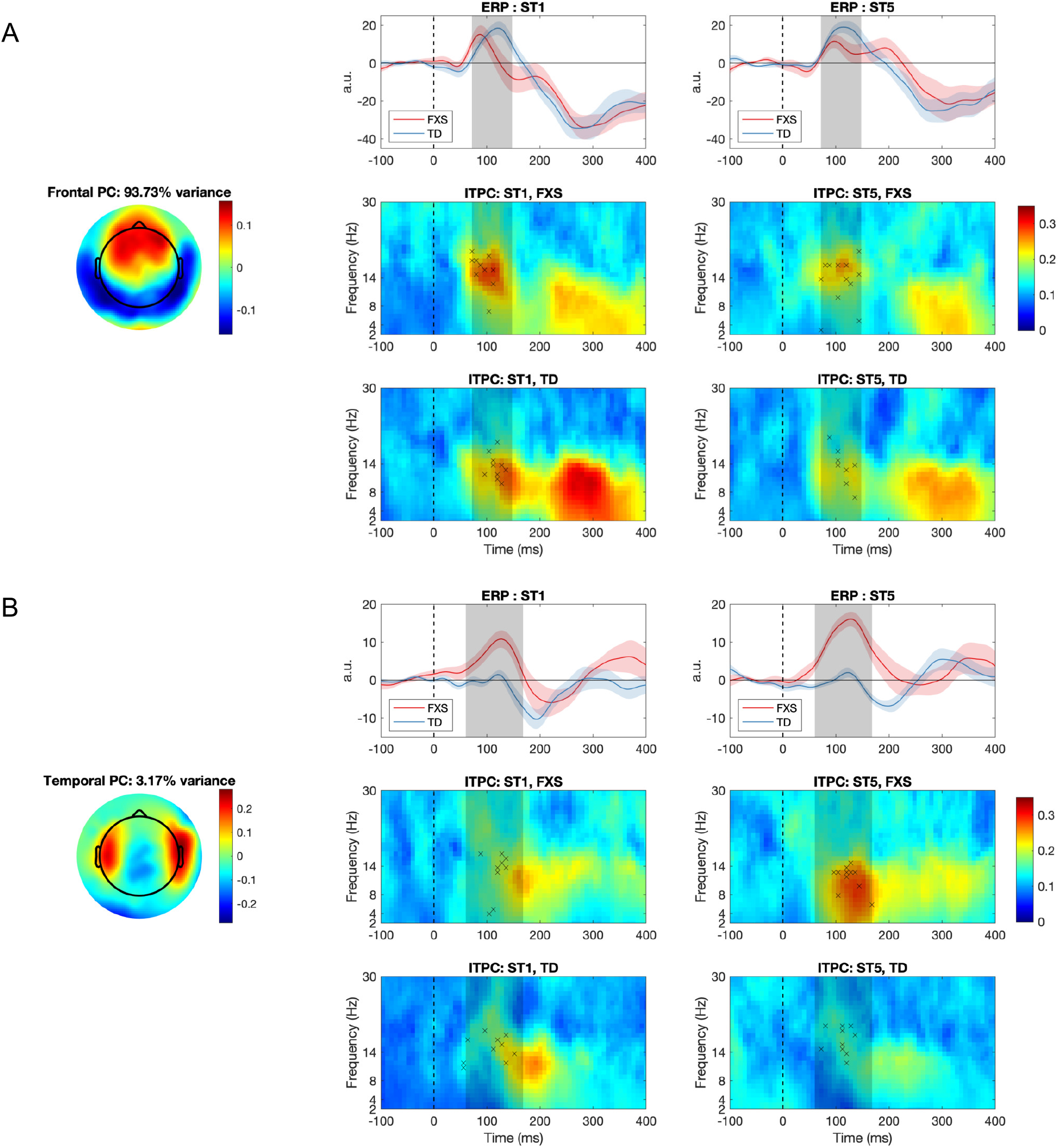
The (A) frontal and (B) temporal principal component (PC) and their associated event-related potential (ERP) and inter-trial phase coherence (ITPC) for the FXS and the TD group. Dashed lines represent the stimulus onset; the red and blue shaded areas denote standard error of the mean; the grey shaded areas denote the P1 window identified by a non-parametric statistical analysis; crosses in the ITPC plots indicate where maximum ITPC was identified for each participant a.u., arbitrary unit

#### 2.4.2 Event-related potentials

The PCs derived from the previous step were used as spatial filters to scale and integrate signals from all channels in each trial, for each condition and each participant. This reduced the 128-channel EEG data to one single time course for each trial. Event-related potential (ERP) analysis was conducted with these PC-transformed time courses instead of the original multi-channel signals.

We specifically focused on the P1 component of an ERP in this study, which is the first positive deflection in the average EEG waveform after the stimulus onset. Considering that the ERP waveform changes during childhood (Wunderlich et al., 2006), and that we are uncertain about the exact latency for P1 in children with FXS, we did not use a pre-defined time window for P1 calculation. Instead, we performed a non-parametric cluster-based permutation test (Maris and Oostenveld, 2007) for each condition and group. This method searched for continuous time intervals in which ERP is statistically above zero at around 100ms, and we defined the P1 window as the union of such intervals over both conditions and both groups. The amplitude of P1 was then calculated as the peak in each participant’s PC-transformed time course within this window. Habituation of P1 was defined as the difference in P1 amplitude between ST1 and ST5 (i.e., ST1 - ST5) as we expected to observe a reduced P1 amplitude in ST5 compared to ST1.

#### 2.4.3 Inter-trial phase coherence

Inter-trial phase coherence (ITPC) examines the consistency in oscillatory phase across all the trials in a condition. It can be calculated with the following equation:

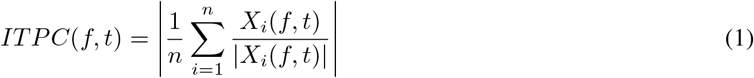

where *X*_*i*_(*f, t*) denotes the Short-Time Fourier Transform (STFT) of a given time course *x*(*t*), and *n* denotes the number of trials in a condition. The value of ITPC ranges between 0 and 1, with 0 indicating totally random phase distribution and 1 indicating perfect phase synchronization.

We applied a STFT with a 32-point Hann window, 95% overlap between windows, and a 256-point fast Fourier Transform. We chose a narrow window to capture transient phase changes at the cost of spectral resolution, because the exact frequency of peak phase coherence is not the interest of this study. After a time-frequency map of ITPC was calculated for one condition, we applied the P1 window derived from the ERP analysis in the previous step, and searched for the maximum ITPC value within that time window and across all the frequencies (2 – 30Hz), which is the ITPC measure we used for further analysis. As in the ERP analysis, habituation of ITPC was defined as the difference in ITPC between ST5 and ST1.

### 2.5 Clinical measures

Receptive and expressive language abilities were evaluated by the Preschool Language Scale - 5th Edition (PLS) (Zimmerman et al., 2011), a comprehensive developmental language assessment standardized for children aged 0 – 83 months. The Vineland Adaptive Behavior Scales - 3rd Edition (VAS) (Sparrow et al., 2018), a parent-report measure assessing communication, social, motor, and daily living skills commonly used in clinical trials, was also administered. Standard scores and v-scaled scores were used for PLS and VAS, respectively. Non-verbal skills were evaluated by the Mullen Scales of Early Learning (MSEL) (Mullen, 1989), a standardized assessment of development for children 0 – 69 months of age; a nonverbal developmental quotient (NVDQ) was calculated for all FXS participants and TD participants under 70 months of age based on their fine motor and visual reception scores. We also included the Child Sensory Profile-2 (SPC) (Dunn, 2014) to evaluate sensory processing patterns. We calculated a customized score for sensory hypersensitivity by summing the raw score of 13 selected questions in the Child Sensory Profile-2 caregiver questionnaire — these questions were picked from the Auditory Processing, Visual Processing and Touch Processing sections, and from the Avoiding and Sensitivity quadrants, which are highly pertinent to the sensory hypersensitivity characteristic that we are interested to study in patients with FXS. The exact questions selected for SPC calculation are listed in Supplementary Material Table S1. Two FXS and two TD participants did not complete the SPC questionnaire, thus were excluded from this part of the analysis. A summary of clinical measures is shown in Table 2.

**Table 2:**
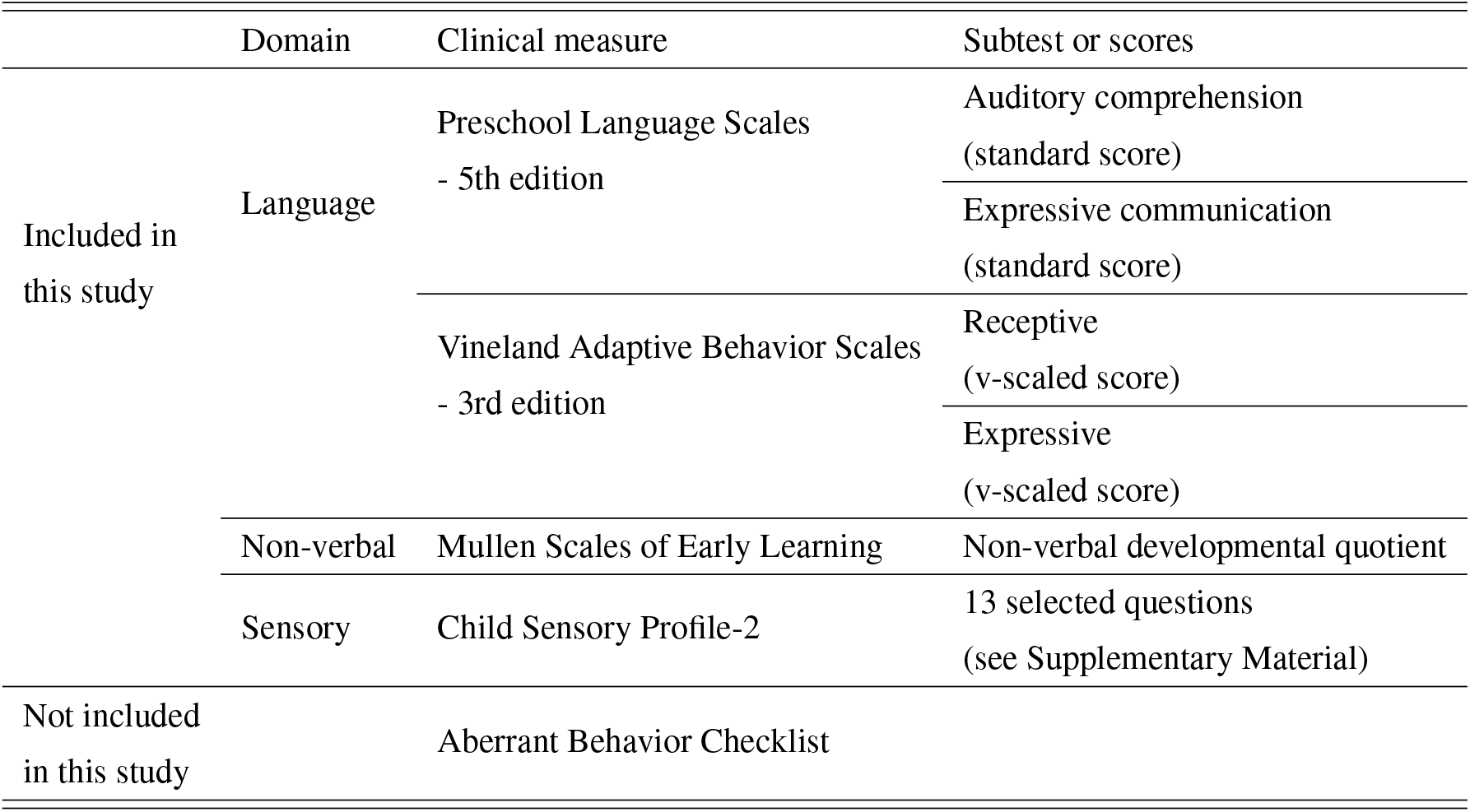
Behavioral data collected from participants

### 2.6 Linear regression analysis

The association between EEG and behavior was explored with a series of linear regression analyses. We used an ordinary least squares (OLS) model to search for a linear relationship between dependent variables (i.e., behavioral measures) and independent variables (i.e., EEG measures) interacted with group identify (i.e. FXS or TD). We also included the age of participants as a covariate to parse out any effect of age on EEG or behavior. The equation for this OLS model (Model 1) is as follows:

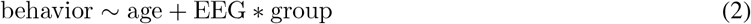

We used the built-in functions in R (R Core Team, 2020) to calculate the beta coefficient of each term in the model. In cases where the interaction term has a significant beta value (p<0.05), i.e., the two groups have different effects, we conducted a marginal effect analysis on the previous OLS model to estimate the effect within each group. If no significant interaction was observed, we discarded the interaction term and estimated the effect with a new OLS model with age and group being covariates (Model 2):

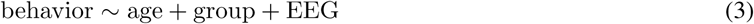

With this general framework for regression analysis, we examined the association between each pair of EEG and behavioral measures. There are four EEG measures in this study: 1) P1 amplitude of ST5, 2) P1 habituation, 3) ITPC of ST5, and 4) ITPC habituation; and there are six clinical measures: 1) auditory comprehension in PLS (PLS-R), 2) expressive communication in PLS (PLS-E), 3) receptive language in VAS (VAS-R), 4) expressive language in VAS (VAS-E), 5) NVDQ in MSEL, and 6) sensory sensitivity and avoiding in SPC. In total, twenty-four models were estimated for each spatial PC to study its clinical correlate.

### 2.7 Statistical analysis

We conducted a series of two-tailed two-sample t-tests to compare the demographics, behavioral scores, and EEG measures within each condition between the FXS and the TD group. Between ST1 and ST5 conditions, we conducted right-tailed paired t-tests on EEG measures within each group.

For the linear regression analysis, we applied the Bonferroni correction to control for family-wise error rate. A factor of 2 was applied to p values in SPC and NVDQ models (i.e., for the two types of EEG measures, P1 and ITPC, analyzed in this study); a factor of 4 was applied to p values in PLS and VAS models (i.e., for the two types of EEG and the two types of language assessments). No correction was applied between EEG amplitude and habituation, or among different domains of clinical measures, as they each formed an independent hypothesis.

## 3 Results

### 3.1 Sample description

Demographic data, including the MSEL non-verbal developmental quotient (NVDQ), PLS and VAS language scores, and Child Sensory Profile-2 (SPC) scores are shown in Table 1. The FXS and the TD groups are age-matched (p = 0.7914), but have substantially different NVDQ, and receptive and expressive language abilities (p<0.0001 for all). The SPC scores are comparable between the two groups (p = 0.9437).

### 3.2 Neural response and habituation

Brain responses to repeated tones within a passive auditory oddball paradigm were analysed using a spatial principal component (PC) analysis. Two significant PCs were identified in the parallel analysis, one with high weights in the frontal region (i.e., the frontal PC; Figure. 3A) and the other with high weights in the temporal regions (i.e., the temporal PC; Figure. 3B). The PC-transformed time courses for the frontal PC show a clear ERP waveform with a P1 peak at around 100ms in both FXS and TD. The average ITPC associated with P1 is stronger in ST1 than in ST5 in both groups. In the temporal PC, however, only the FXS group shows a strong P1 response in both ST1 and ST5; the TD group does not show a clear P1 peak within the P1 window.

The P1 amplitude in the frontal PC is comparable between FXS and TD in both the ST1 (p = 0.7593, Cohen’d = 0.127) and the ST5 (p = 0.4190, Cohen’s d = 0.337) conditions (Figure 4A). No significant difference in P1 amplitude between ST1 and ST5 (i.e., the habituation effect) was identified in either FXS (p = 0.4386, Cohen’s d = 0.046) or TD (p = 0.6293, Cohen’s d = 0.107) (Figure 4A). The ITPC is also similar between groups in both conditions in the frontal PC (Figure 4B). However, we observed habituation effects in ITPC in both the FXS (p = 0.0149, Cohen’d = 0.711) and the TD (p = 0.0244, Cohen’s d = 0.709) group (Figure 4B).

**Figure 4:**
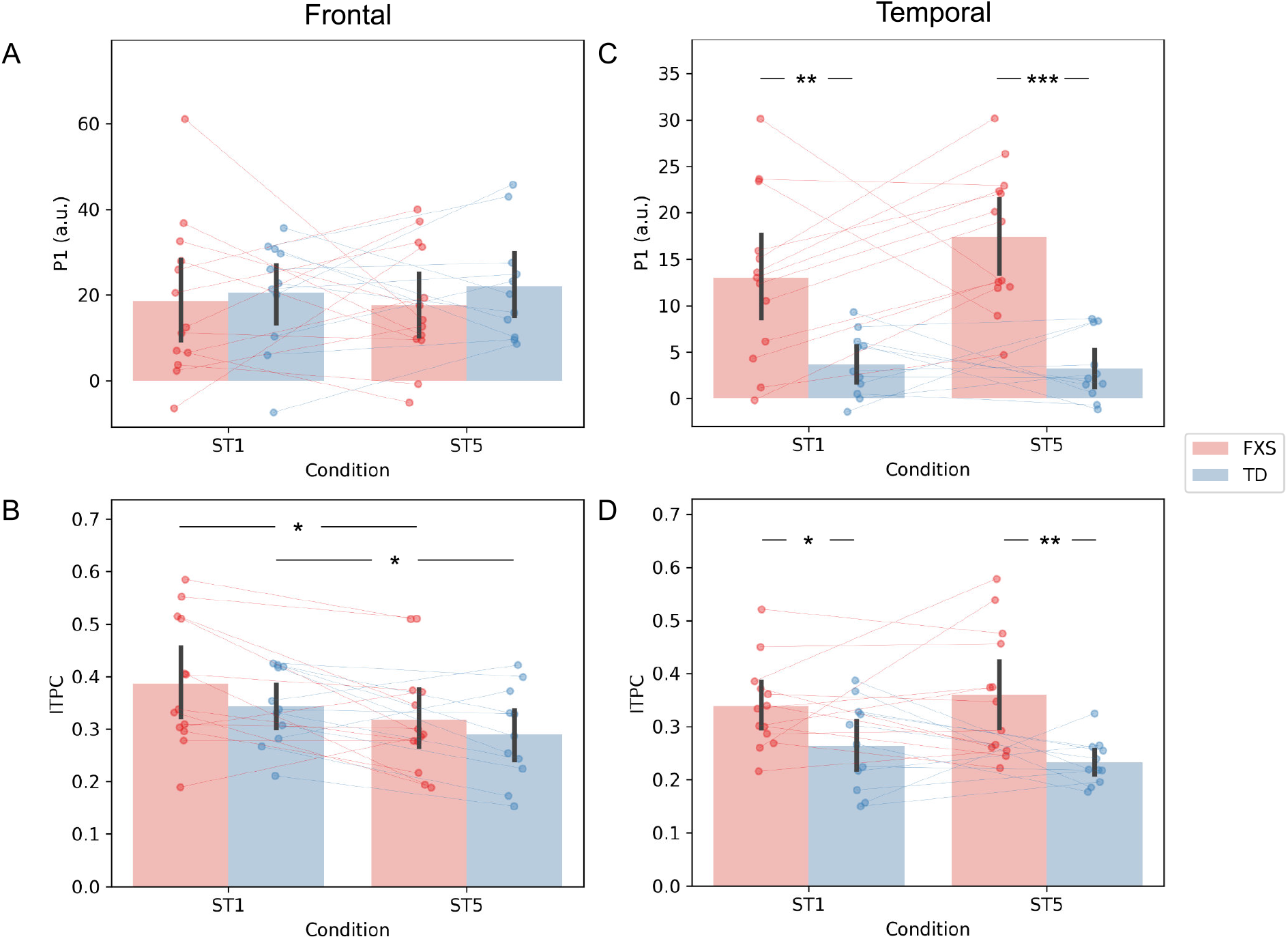
Comparison of (A) frontal P1, (B) frontal ITPC, (C) temporal P1, and (D) temporal ITPC between conditions and groups *p<0.05; **p<0.01; ***p<0.001

The EEG measures in the temporal PC show a contrasting pattern than those in the frontal PC. The FXS group exhibit a more prominent P1 amplitude than the TD group in both the ST1 (p = 0.0037, Cohen’s d = 1.328) and the ST5 (p < 0.0001, Cohen’s d = 2.357) conditions (Figure 4C). Their ITPC measures are also significantly different in ST1 (p = 0.0402, Cohen’s d = 0.893) and ST5 (p = 0.0027, Cohen’s d = 1.383) (Figure 4D). No significant habituation effects were observed in this PC.

### 3.3 Relationship between EEG and behavior

We examined the relationship between EEG and clinical measures through a series of linear regression analyses. Table 3 shows the results of linear regression analyses where an interaction term for EEG and group identify were included (Model 1), along with age as a covariate. Further analyses of significant interactions are described below. For regressions where the interaction term was not significant, a simplified regression (Model 2) was performed (Supplementary Material Table S2); however, no significant associations between EEG and clinical measures were observed.

**Table 3:**
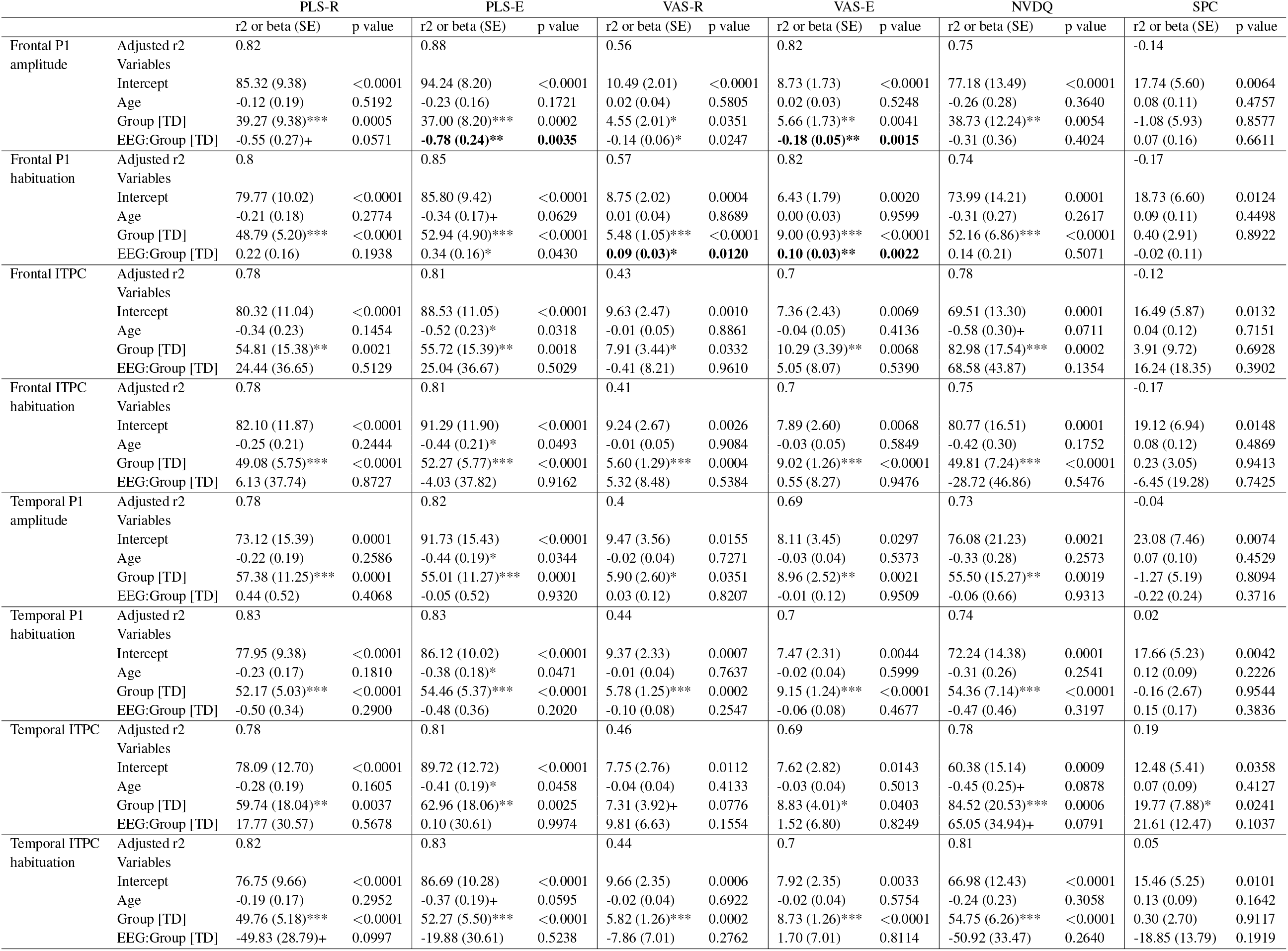
Regression analysis results for the ordinary least squares model with an interaction term between EEG and group identify. Significant p values for the interaction term that survived the Bonferroni correction are in bold. +p< 0.1; *p<0.05; **p<0.01; ***p<0.001

#### 3.3.1 EEG and language

The models for language scores (i.e., PLS-R, PLS-E, VAS-R and VAS-E) generally show significant or marginally significant interactions between P1 measures (i.e., P1 amplitude of ST5 and P1 habituation) and group identity (Table 3), indicating that the two groups may have different P1-behavior relationships. Accordingly, we analyzed the marginal effects of the model for FXS and TD separately to reveal such group-level differences. Results show that the frontal P1 amplitude of ST5 is negatively correlated with all language scores only in the FXS group, among which its relationship with PLS-E and VAS-E survived correction for multiple comparisons (Table 4). Additionally, the frontal P1 habituation showed strong, positive association with VAS-R (p = 0.0055) and VAS-E (p = 0.0004), also only in FXS. These linear relationships are depicted in Figure 5 and 6.

**Table 4:** Marginal effect results for the ordinary least squares model with an interaction term. Significant p values that survived the Bonferroni correction are in bold. *p<0.05; **p<0.01; ***p<0.001

|  |  | PLS-R |  | PLS-E |  | VAS-R |  | VAS-E |  |
| --- | --- | --- | --- | --- | --- | --- | --- | --- | --- |
|  |  | dy/dx | p value | dy/dx | p value | dy/dx | p value | dy/dx | p value |
| Frontal P1 amplitude | FXS | -0.55 (0.27)* | 0.0428 | **-0.78 (0.24)\*\*\*** | **0.0009** | -0.14 (0.06)* | 0.0147 | **-0.18 (0.05)\*\*\*** | **0.0002** |
|  | TD | 0.00 (0.30) | 0.9954 | 0.10 (0.26) | 0.7104 | -0.07 (0.06) | 0.2623 | 0.00 (0.06) | 0.9587 |
| Frontal P1 habituation | FXS | 0.22 (0.16) | 0.1779 | 0.34 (0.16)* | 0.0301 | **0.09 (0.03)\*\*** | **0.0055** | **0.10 (0.03)\*\*\*** | **0.0004** |
|  | TD | -0.13 (0.27) | 0.6423 | -0.10 (0.26) | 0.6972 | -0.01 (0.05) | 0.8587 | -0.02 (0.05) | 0.6178 |

**Figure 5:**
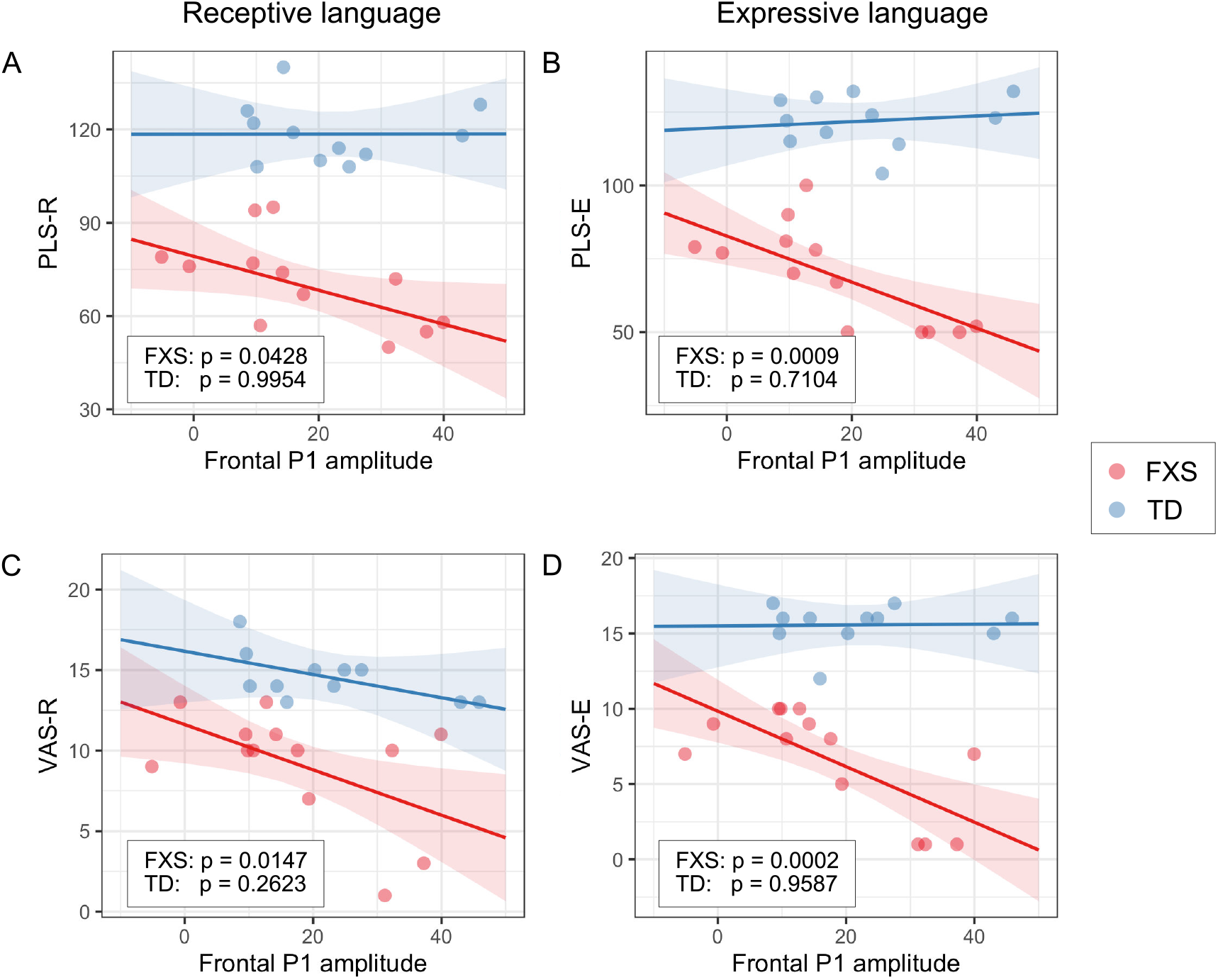
Association between the frontal P1 amplitude of ST5 and (A) PLS-R, (B) PLS-E, (C) VAS-R, and (D) VAS-E in Model 1. The lines and shaded areas denote the prediction lines and their 95% confidence interval estimated by the marginal effect. The scattered dots represent individual data. The p value of marginal effect for each group is shown in the legend.

**Figure 6:**
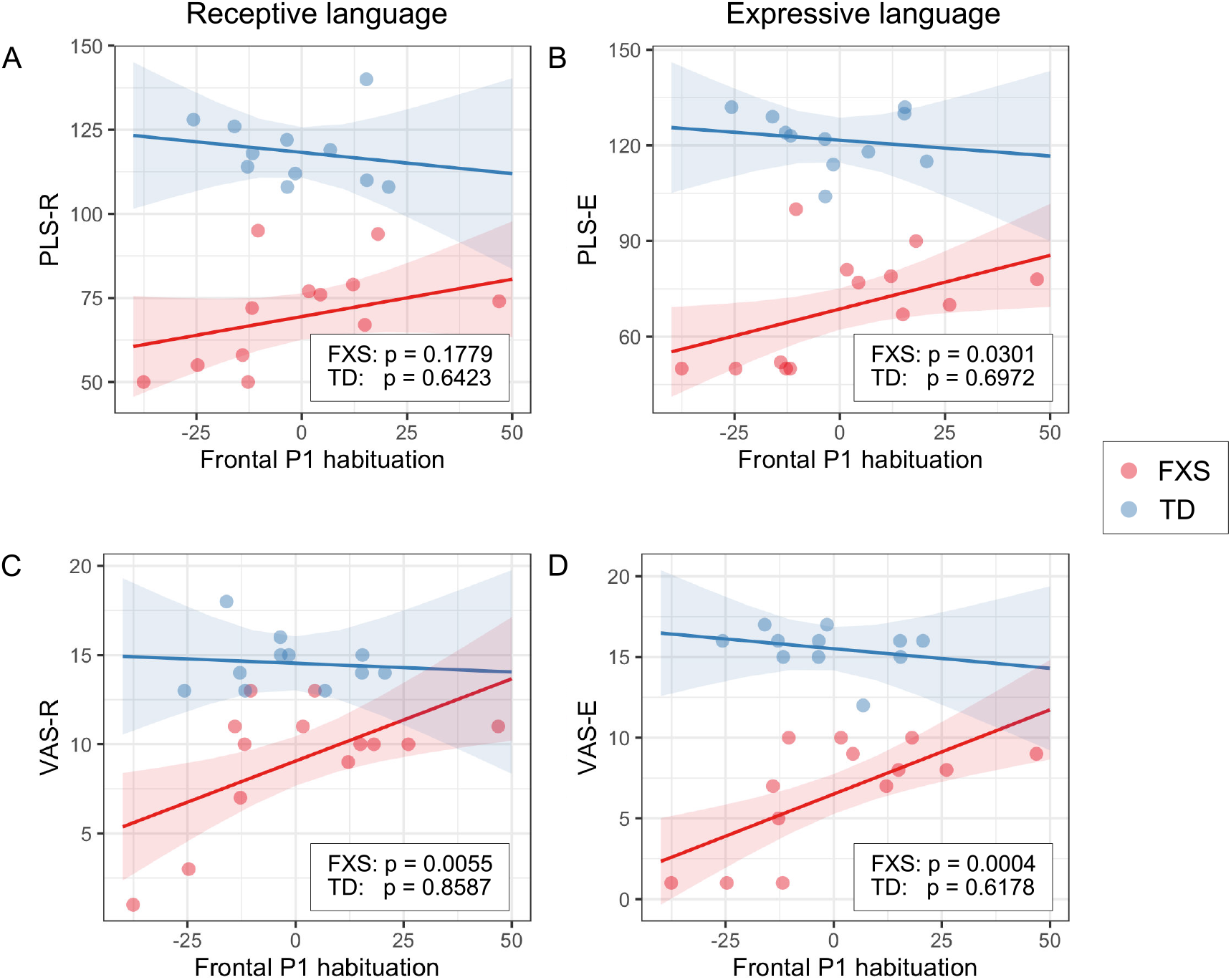
Association between the frontal P1 habituation and (A) PLS-R, (B) PLS-E, (C) VAS-R, and (D) VAS-E in Model 1. The lines and shaded areas denote the prediction lines and their 95% confidence interval estimated by the marginal effect. The scattered dots represent individual data. The p value of marginal effect for each group is shown in the legend.

#### 3.3.2 EEG and non-verbal skills / sensory hypersensitivity

We also examined the relationship between EEG and NVDQ or SPC score. Neither Model 1 nor Model 2 revealed significant association between NVDQ / SPC score and any of the EEG measures. Only the temporal ITPC was found to be marginally correlated with NVDQ before correction for multiple comparisons (p = 0.0791, Table 3). These findings suggest that the strong association with EEG discovered in Section 3.3.1 may be specific to language development.

## 4 Discussion

In this study, we compared the amplitude and habituation of the auditory P1 response and its corresponding inter-trial phase coherence (ITPC) between male children with and without FXS, after performing a spatial principal component analysis (PCA) on their EEG data. We also examined the association between these EEG measures and several clinical measures that assessed sensory sensitivities, language abilities, and non-verbal development. The results show that though the two groups exhibit comparable P1 amplitude and ITPC in the frontal PC, male children with FXS have stronger temporal P1 and ITPC. No difference in habituation pattern was found between the two groups. In terms of the clinical correlate, we discovered a strong association between language abilities and the amplitude and habituation of the frontal P1 in individuals with FXS.

### 4.1 Interpretation of PCA results

The spatial PCA identified two significant PCs in our EEG data — one with high weights in the frontal region, and the other with high weights in the lateralized temporal regions (Figure 3). The heightened P1 amplitude observed in the temporal PC in the FXS group can be understood in two ways. First, elevated N1 and P2 responses have previously been reported as a robust EEG phenotype in individuals with FXS (Clair et al., 1987; Castrén et al., 2003; Knoth et al., 2014; Ethridge et al., 2016), and both have been linked to auditory hypersensitivity and auditory processing alterations in FXS (Schneider et al., 2013; Rotschafer and Razak, 2014). Therefore, the strong temporal P1 response observed in FXS in this study could possibly be caused by impaired inhibition in the auditory cortex. Second, the topographical location of peak N1 has been shown to change with development; prior to the age of six it is largely temporal (Bruneau et al., 1997) and then this shifts to central regions in older ages (Tonnquist-Uhlén et al., 1995; Knoth and Lippé, 2012). Given the adjacency of P1 and N1 sources in the auditory cortex (Yvert et al., 2005), we expect a similar temporal-to-central developmental trajectory in the topographical pattern of P1 in children. With this hypothesis, the findings in this study might have reflected a delayed development of the auditory brain in children with FXS, which is part of the global developmental delay known in this population (Roberts et al., 2016; Wheeler et al., 2021). Either interpretation points to the fact that children with FXS may have altered (impaired or delayed) auditory processing than their age-matched TD peers.

### 4.2 ERP and ITPC differences between groups

Different from our hypothesis, no habituation of P1 amplitude was observed in either group in this study (Figure 4A & C). Previous research adopted a similar experimental setup using repeated auditory stimuli, and reported habituation of N1 in the control group, but not in FXS patients (Castrén et al., 2003; der Molen et al., 2012a). However, the participants in these studies were either adults or older children (7 - 13 years) compared to our cohort. Given that the waveform of auditory ERP changes throughout childhood (Wunderlich et al., 2006), the results from these studies may not be directly transferable to the current one. More importantly, despite that N1 and P1 share many commonalities, they are substantially different in their sensitivity to stimulus presentation rate. According to the neural adaptation theory (Kudela et al., 2018), both would reduce their intensity if a train of stimuli were played. However, P1 would recover to its full amplitude if the stimuli are apart by as short as a few hundred milliseconds (Picton, 2010), while the recovery of N1 takes more than 10 seconds (Cowan et al., 1993). Since the inter-stimulus interval (ITI) was set at 1000ms in this experiment, there was sufficient time for P1 to return to its baseline amplitude, which might have explained the absence of P1 habituation in here. Future studies with a shorter ITI (a few hundred milliseconds) are warranted.

In spite of the null findings in P1 amplitude habituation, we observed strong habituation of frontal ITPC in both groups (Figure 4B). Biomedical signals, like EEG, have three important characteristics — amplitude, frequency, and phase. Even though event-related potential (ERP) is a popular choice of neural signature for studying human brains, it mainly accounts for the “amplitude” aspect of a signal. ITPC, on the other hand, utilizes time-frequency decomposition to examine synchrony of instantaneous phase across trials at different frequencies, which discards the magnitude information through normalization (see Equation 1) and focuses more on “phase” and “frequency”. Therefore, it may provide information that is not available in the aggregate ERP waveform (Makeig et al., 2004). Additionally, ITPC was found to be a more stable neural marker than evoked spectral power in terms of inter-subject variability (Engel et al., 2020) and test-retest reliability (Legget et al., 2017), possibly because it is independent of amplitude (Legget et al., 2017). In developmental neuroscience, ITPC is relatively less studied than ERP, yet previous studies report weaker gamma ITPC (Ethridge et al., 2016, 2019) and reduced habituation of low-frequency (theta-alpha band) ITPC (Ethridge et al., 2016) in individuals with FXS compared to those without FXS, demonstrating the clinical relevance of ITPC measures as a potential biomarker in FXS research. Here, we report habituation of frontal ITPC in both FXS and TD groups (Figure 4B), and heightened temporal ITPC in FXS compared to TD (Figure 4D). Cortical phase synchrony is considered to be modulated by cognitive demands (Nash-Kille and Sharma, 2014), and it has been associated with various cognitive processes like information processing (Tass et al., 1998), feature binding (Palva et al., 2005), and neural computation (Fries, 2005). Therefore, the results in this study suggest a relatively lower cognitive demand for information processing after a few repetitions of the same standard stimulus, a mechanism that is shared by, but also altered in boys with FXS. More future studies on ITPC in FXS children are needed to confirm these findings.

### 4.3 Clinical correlations of brain measures

We observed a pronounced linear relationship between language scores and P1 amplitude (Figure 5) and habituation (Figure 6) in the FXS group; weaker P1 response to late standard stimuli (i.e., ST5) as well as stronger habituation of P1 is associated with higher receptive and expressive language abilities. At the time when ST5 is played, the same standard stimulus has been repeated for four times. The level of information novelty in ST5 is extremely low. Therefore, it is cognitively advantageous that our neural system reacts weakly to yet another standard stimulus, so that neural resources can be preserved for other cognitive processes. Huber and O’Reilly (2003) proposed a short-term synaptic depression model to explain this phenomenon. In this model, the response to a recently identified object is suppressed, while any new object, for its high salience, triggers stronger neural activities. This mechanism of neural habituation was considered to aid perceptual processing of a novel object (Huber and O’Reilly, 2003). Later, an EEG study by Jacob and Huber (2020) confirmed the benefit of this habituation mechanism in working memory and novelty detection. Furthermore, a recent behavioral study by Marino and Gervain (2019) linked the novelty detection ability of infants measured at 9 months with their future language outcomes at 12, 14, 18, and 24 months. These previous works suggest a beneficial role of neural habituation in language learning, laying the foundation for understanding our findings in the FXS group. In the TD group, however, we did not observe a strong impact of habituation on language scores. This result could be interpreted from two angles. First, the range of language scores in the TD group is much narrower than that in the FXS group. Thus, it is mathematically more difficult to achieve significance in linear regression models from the TD group, especially given our small sample size (da Silva and Seixas, 2017). Second, the children in the TD group have an intact, unimpaired neural system to support their early language learning. After the critical language period, further language development in these participants may become less sensitive to the cognitive advantages gained from neural habituation, but rather be driven by other aspects of learning such as language exposure, home support, or non-verbal skills. However, for individuals still in early language acquisition and development (like our study’s FXS group), auditory habituation may play a more important role.

The neural markers we examined in this study have the potential to become reliable biomarkers for early diagnosis and outcome prediction in FXS. In a recent review, Kenny et al. (2022) summarized the EEG studies on FXS in the past decades, and suggested a few EEG features, including the N1 amplitude, gamma oscillatory power, and gamma phase-locking, as promising translational biomarkers for evaluating the efficacy of FXS treatment. Here, we add the P1 amplitude and habituation to this list of promising candidates. The passive auditory oddball paradigm is excellent for studying neurodevelopment in children or infants, as it requires no attention or reaction from the participants. The repeated standard stimuli used in this paradigm can elicit differential neural response in the FXS and the TD group, which can be effectively captured by an unsupervised learning algorithm like PCA. The pronounced P1 amplitude in the temporal regions observed in our participants with FXS could be employed as a marker for early diagnosis of this syndrome, if the same pattern can be found in infants. Similarly, P1 amplitude and habituation have the potential to be markers for predicting language outcomes in young patients with FXS, which will facilitate the implementation of early intervention and therapeutics for these patients. They may also serve as an outcome measure for clinical trials to evaluate the efficacy of treatment. Futures studies on a younger population are therefore desirable.

### 4.4 Limitations

This study is bound to certain limitations. The sample size in this study is small compared to other EEG studies with adults. Collecting data, especially high-quality EEG data, from children with FXS can be challenging, as they often are sensitive to being touched (especially on their heads), have limited expressive language, and have difficulty sitting in one place. We were successful in obtaining high quality EEG data in 80% of the FXS participants eligible for this study (12/15). To do this, we communicated with parents ahead of visits in order to implement participant-specific behavioral strategies, visual schedules, and positive reinforcements. Careful considerations should always be made to accommodate the needs of these participants, which usually takes a long time for training and accumulating experience. Hence we expect studies with much larger sample sizes to happen through multi-site collaborative research. Another limitation of this study was the inclusion of only full-mutation males with FXS. Understanding the neurobiology of females with FXS is important, as the mosaic expression of FMRP in this group may allow for improved understanding of the role of FMRP in neurodevelopment and provide insight into therapeutic strategies focused on genetic therapy. However, given the increased heterogeneity within females with FXS, larger samples will be needed.

## 5 Conclusion

We analyzed the auditory evoked response to repeated sounds in male children with or without FXS, and examined the neural correlate to early language development. The P1 amplitude and inter-trial phase coherence in the temporal regions were found to be increased in individuals with FXS compared to their age-matched typically developing peers. Additionally, the P1 amplitude and habituation in the frontal region were strongly associated with language scores in the FXS group. These findings suggest that the auditory P1 might be a potential biomarker for early diagnosis of FXS, and for predicting language outcomes in children with this syndrome.

## Data Availability

Data produced in the present study are available upon reasonable request to the authors.

## Author Contributions

CAN and CLW contributed to conception and design of the study. CLW recruited participants and collected the data. WWA performed the data analysis and data visualization. WWA and CLW wrote the draft of the manuscript. All authors contributed to manuscript revision, read, and approved the submitted version.

## Acknowledgments

We thank all the families who participated in this study. We also thank Jack Keller, Megan Lauzé, John Fitzgerald, Megan Hartney and the Translational Neuroscience Center Human Neurobehavioral Core at Boston Children’s Hospital for their assistance in data collection of FXS and age-matched controls. We also thank Graham Holt for his EEG technical support.

## Supplemental Data

Supplementary materials are available for this manuscript.

## Data Availability Statement

The data sets used and/or analyzed during the current study are available from the corresponding author upon reasonable request.

