## Supplemental Tables for "Neural response to repeated auditory stimuli and its association with early language development in children with Fragile X syndrome"

### SUPPLEMENTARY MATERIAL

A medRχiv PREPRINT

July 5, 2022

| Sensory | Quarant | Question |
|---|---|---|
| Auditory | Avoiding | Reacts strongly to unexpected or loud noises (for example, sirens, dog barking, hair dryer) |
| | | Holds hands over ears to protect them from sound |
| | | Becomes unproductive with background noise (for example, fan, refrigerator) |
| | Sensitivity | Struggles to complete tasks when music or TV is on |
| | | Is distracted when there is a lot of noise around |
| | | Tunes me out or seems to ignore me |
| | | Seems not to hear when I call his or her name (even though hearing is OK) |
| Visual | Sensitivity | Prefers to play or work in low lighting |
| | | Is more bothered by bright lights than other same-aged children |
| Touch | Avoiding | Shows an emotional or aggressive response to being touched |
| | Sensitivity | Shows distress during grooming (for example, fights or cries during haircutting, face washing, fingernail cutting) |
| | | Becomes anxious when standing close to others (for example, in a line) |
| | | Rubs or scratches a part of the body that has been touched |

Table 1: Questions selected from the Sensory Profile Child questionnaire for calculating the SPC score

| | | PLS-R | | PLS-E | | VAS-R | | VAS-E | | NVDQ | | SPC | |
|---|---|---|---|---|---|---|---|---|---|---|---|---|---|
| | | r2 or beta (SE) | p value | r2 or beta (SE) | p value | r2 or beta (SE) | p value | r2 or beta (SE) | p value | r2 or beta (SE) | p value | r2 or beta (SE) | p value |
| Frontal P1 amplitude | Adjusted r2 | Not analyzed | | Not analyzed | | Not analyzed | | Not analyzed | | 0.74 | | -0.07 | |
| | Variables | in Model 2 | | in Model 2 | | in Model 2 | | in Model 2 | | | | | |
| | Intercept | | | | | | | | | 76.70 (13.75) | 0 | 17.70 (5.43) | 0.0049 |
| | Age | | | | | | | | | -0.36 (0.27) | 0.2028 | 0.07 (0.10) | 0.4732 |
| | Group | | | | | | | | | 52.16 (7.03)*** | 0 | -0.19 (2.87) | 0.947 |
| | EEG | | | | | | | | | 0.01 (0.27) | 0.9679 | 0.09 (0.12) | 0.4461 |
| Frontal P1 habituation | Adjusted r2 | Not analyzed | | Not analyzed | | Not analyzed | | Not analyzed | | 0.74 | | -0.1 | |
| | Variables | in Model 2 | | in Model 2 | | in Model 2 | | in Model 2 | | | | | |
| | Intercept | | | | | | | | | 76.29 (14.12) | 0 | 18.95 (6.22) | 0.0077 |
| | Age | | | | | | | | | -0.35 (0.26) | 0.1994 | 0.08 (0.11) | 0.4423 |
| | Group | | | | | | | | | 52.31 (6.89)*** | 0 | 0.35 (2.80) | 0.9023 |
| | EEG | | | | | | | | | 0.02 (0.18) | 0.9013 | -0.03 (0.08) | 0.6969 |
| Frontal ITPC | Adjusted r2 | 0.79 | | 0.82 | | 0.43 | | 0.71 | | 0.74 | | -0.05 | |
| | Variables | | | | | | | | | | | | |
| | Intercept | 81.40 (11.21) | 0 | 87.75 (11.17) | 0 | 10.55 (2.54) | 0.0005 | 7.39 (2.46) | 0.007 | 76.84 (14.80) | 0.0001 | 15.86 (5.79) | 0.0145 |
| | Age | -0.30 (0.21) | 0.1705 | -0.49 (0.21)* | 0.0291 | -0.01 (0.05) | 0.8479 | -0.04 (0.05) | 0.4277 | -0.35 (0.30) | 0.2479 | 0.04 (0.11) | 0.7057 |
| | Group | 48.90 (5.36)*** | 0 | 53.04 (5.34)*** | 0 | 5.29 (1.21)*** | 0.0003 | 8.97 (1.17)*** | 0 | 52.21 (6.88)*** | 0 | 0.30 (2.71) | 0.9139 |
| | EEG | 10.17 (32.38) | 0.7567 | 18.48 (32.26) | 0.5731 | -2.91 (7.34) | 0.6958 | 3.49 (7.10) | 0.6284 | -1.12 (40.78) | 0.9784 | 16.24 (17.78) | 0.3745 |
| Frontal ITPC habituation | Adjusted r2 | 0.79 | | 0.82 | | 0.47 | | 0.71 | | 0.79 | | -0.11 | |
| | Variables | | | | | | | | | | | | |
| | Intercept | 81.70 (10.37) | 0 | 89.68 (10.41) | 0 | 9.22 (2.27) | 0.0006 | 7.95 (2.29) | 0.0024 | 67.78 (13.13) | 0.0001 | 17.83 (6.17) | 0.0107 |
| | Age | -0.26 (0.18) | 0.1756 | -0.43 (0.18)* | 0.0315 | -0.01 (0.04) | 0.7484 | -0.03 (0.04) | 0.5251 | -0.28 (0.23) | 0.2507 | 0.10 (0.10) | 0.3185 |
| | Group | 48.87 (5.31)*** | 0 | 52.77 (5.33)*** | 0 | 5.50 (1.16)*** | 0.0001 | 8.89 (1.17)*** | 0 | 53.91 (6.24)*** | 0 | 0.53 (2.85) | 0.8539 |
| | EEG | 13.49 (30.67) | 0.6649 | 10.47 (30.79) | 0.7373 | 8.84 (6.71) | 0.2024 | -0.06 (6.78) | 0.9935 | 72.53 (34.88)+ | 0.0513 | -0.17 (16.25) | 0.9917 |
| Temporal P1 amplitude | Adjusted r2 | 0.79 | | 0.82 | | 0.43 | | 0.71 | | 0.74 | | -0.01 | |
| | Variables | | | | | | | | | | | | |
| | Intercept | 75.93 (14.37) | 0 | 95.36 (14.49) | 0 | 9.63 (3.29) | 0.0084 | 8.32 (3.20) | 0.0169 | 81.92 (18.75) | 0.0003 | 24.07 (7.21) | 0.0042 |
| | Age | -0.24 (0.19) | 0.2236 | -0.45 (0.19)* | 0.0262 | -0.02 (0.04) | 0.7066 | -0.03 (0.04) | 0.5106 | -0.38 (0.26) | 0.1625 | 0.07 (0.09) | 0.4422 |
| | Group | 53.21 (8.52)*** | 0 | 49.62 (8.59)*** | 0 | 5.65 (1.95)** | 0.009 | 8.64 (1.89)*** | 0.0002 | 48.84 (10.74)*** | 0.0002 | -3.45 (4.10) | 0.4123 |
| | EEG | 0.32 (0.47) | 0.5053 | -0.21 (0.47) | 0.663 | 0.02 (0.11) | 0.8512 | -0.02 (0.10) | 0.8718 | -0.24 (0.58) | 0.688 | -0.28 (0.22) | 0.2217 |
| Temporal P1 habituation | Adjusted r2 | 0.82 | | 0.82 | | 0.45 | | 0.71 | | 0.75 | | 0.01 | |
| | Variables | | | | | | | | | | | | |
| | Intercept | 80.66 (9.13) | 0 | 89.79 (9.94) | 0 | 9.82 (2.23) | 0.0003 | 7.79 (2.19) | 0.002 | 75.71 (13.63) | 0 | 18.32 (5.23) | 0.0029 |
| | Age | -0.27 (0.17) | 0.1171 | -0.44 (0.18)* | 0.0263 | -0.02 (0.04) | 0.6429 | -0.03 (0.04) | 0.5128 | -0.36 (0.25) | 0.1668 | 0.11 (0.09) | 0.2542 |
| | Group | 51.65 (5.04)*** | 0 | 53.76 (5.49)*** | 0 | 5.70 (1.23)*** | 0.0002 | 9.09 (1.21)*** | 0 | 53.62 (7.02)*** | 0 | -0.29 (2.68) | 0.9144 |
| | EEG | -0.62 (0.30)+ | 0.0528 | -0.24 (0.33) | 0.4737 | -0.07 (0.07) | 0.3562 | -0.04 (0.07) | 0.5761 | -0.30 (0.41) | 0.47 | 0.22 (0.16) | 0.1807 |
| Temporal ITPC | Adjusted r2 | 0.79 | | 0.82 | | 0.51 | | 0.71 | | 0.78 | | -0.01 | |
| | Variables | | | | | | | | | | | | |
| | Intercept | 79.66 (13.11) | 0 | 93.23 (13.16) | 0 | 6.88 (2.78) | 0.0222 | 6.85 (2.87) | 0.027 | 60.59 (15.52) | 0.001 | 13.37 (6.32) | 0.0503 |
| | Age | -0.29 (0.19) | 0.1476 | -0.42 (0.19)* | 0.0406 | -0.04 (0.04) | 0.3602 | -0.03 (0.04) | 0.4398 | -0.51 (0.25)+ | 0.0583 | 0.06 (0.10) | 0.5198 |
| | Group | 50.18 (6.56)*** | 0 | 51.46 (6.58)*** | 0 | 6.81 (1.39)*** | 0.0001 | 9.38 (1.44)*** | 0 | 59.77 (7.59)*** | 0 | 2.84 (3.20) | 0.3881 |
| | EEG | 12.18 (30.98) | 0.6984 | -9.17 (31.08) | 0.771 | 11.61 (6.56)+ | 0.0922 | 3.95 (6.78) | 0.5668 | 65.43 (36.23)+ | 0.0868 | 18.17 (14.28) | 0.2215 |
| Temporal ITPC habituation | Adjusted r2 | 0.79 | | 0.82 | | 0.48 | | 0.71 | | 0.76 | | -0.1 | |
| | Variables | | | | | | | | | | | | |
| | Intercept | 83.68 (10.03) | 0 | 89.96 (10.04) | 0 | 10.53 (2.18) | 0.0001 | 7.87 (2.22) | 0.002 | 78.32 (13.41) | 0 | 18.13 (5.59) | 0.0051 |
| | Age | -0.28 (0.19) | 0.1443 | -0.41 (0.19)* | 0.0396 | -0.03 (0.04) | 0.4603 | -0.02 (0.04) | 0.5645 | -0.41 (0.25) | 0.1247 | 0.10 (0.10) | 0.349 |
| | Group | 49.27 (5.46)*** | 0 | 51.92 (5.46)*** | 0 | 5.77 (1.19)*** | 0.0001 | 8.82 (1.21)*** | 0 | 54.15 (6.86)*** | 0 | 0.76 (2.83) | 0.7913 |
| | EEG | -11.99 (26.52) | 0.6559 | 13.50 (26.53) | 0.6164 | -8.07 (5.77) | 0.1776 | 1.33 (5.86) | 0.8231 | -35.65 (32.01) | 0.2793 | -4.39 (12.89) | 0.7382 |

Table 2: Regression analysis results for the ordinary least squares model without an interaction term (i.e., Model 2). The EEG term in all models are insignificant even before correction for multiple comparison.
+p< 0.1; *p<0.05; **p<0.01; ***p<0.001
